# Genomize-SEQ: An NGS data analysis platform for genomic variant classification and prioritization

**DOI:** 10.1101/2025.09.05.25335160

**Authors:** Ersen Kavak, Tolga Aslan, Ruchan Karaman, Cagatay Aydin, Tolgahan Ozer, Deniz Sunnetci Akkoyunlu, Hakan Savli, Naci Cine, Tuncay Seker

**Affiliations:** Genomize Bilisim ve Biyoteknoloji Anonim Sirketi, 34470, Istanbul/Turkiye; Department of Molecular Biology and Genetics, Faculty of Arts and Sciences, Bogazici University, 34342, Istanbul/Turkiye; Department of Medical Genetics, Faculty of Medicine, Kocaeli University, 41001 Kocaeli/Turkiye

## Abstract

Accurate interpretation of diverse genetic variants remains a pivotal challenge in the diagnosis of rare diseases. Although evidence-based guidelines established by the American College of Medical Genetics and Genomics have enhanced the precision of variant assessment, the practical implementation of this evidence-based classification can be challenging. The inherent genetic heterogeneity in rare diseases, coupled with the need to integrate information from numerous databases, contributes to this complexity. Therefore, advancements in secondary variant calling, automated variant annotation and prioritization, visualization of variant annotations with the raw data, and a streamlined reporting process are crucial for efficient and robust analysis. Here we present Genomize-SEQ, a web-based clinical genomics analysis software that has all of these capabilities, with which more than 300,000 patients have been analyzed to date. Genomize-SEQ collects data from more than 120 different databases to annotate the variants according to ACMG/AMP guidelines and prioritize the variants that could be causative for the clinical presentation of a patient. Genomize-SEQ can also perform real-time data aggregation to calculate variant frequencies in each center as well as the community. This capability helps clinicians to analyze variants more easily in regions without genome projects or in populations underrepresented in existing databases. We validated the annotation capacity of Genomize-SEQ by performing a systematic comparison of ACMG pathogenicity prediction from widely used algorithms and Genomize-SEQ’s algorithm, using ClinGen’s expert curation dataset as a truth set. In addition, we tested the prioritization efficiency of Genomize-SEQ by using real-world whole-exome sequencing data of 215 patients with pre-diagnostic and phenotypic information. Genomize-SEQ identified the causative variants with a 97% success rate, with 52% of these variants ranked in the top position and over 90% ranked within the top 20. Thus, Genomize-SEQ provides a complete solution for comprehensive variant interpretation to achieve fast and reliable diagnosis for rare diseases from next-generation sequencing data.

## Introduction

With recent advances in sequencing technology and substantial decreases in sequencing costs, next-generation sequencing (NGS)-based tests, such as whole genome sequencing (WGS) and whole exome sequencing (WES), are becoming routine clinical practices for both rare disease and cancer diagnosis (Berger and Mardis 2018; Ewans et al., 2022; Sullivan et al., 2023; Brlek et al., 2024). With these tests, thousands of genomic alterations can be detected (Yang et al., 2013; Gargis et al., 2015; Bertoldi et al., 2017). The primary goal of these comprehensive tests is to identify causative variations for diseases with a genetic etiology (Strianese et al., 2020; Vinksel et al., 2021).

The pathogenicity of each variant is assessed by geneticists with the help of bioinformatic tools, based on the set of rules and guidelines defined by the American College of Medical Genetics and Genomics (ACMG) and the Association for Molecular Pathology (AMP) (Richards et al. 2015). Following these guidelines, a total of 28 evidence codes are evaluated and, if applicable, assigned to the variant. Based on the assigned evidence codes and the algorithm outlined in the guidelines, each variant is classified into one of five categories: pathogenic (P), likely pathogenic (LP), variant of uncertain significance (VUS), likely benign (LB), or benign (B) (Richards et al. 2015). Although the ACMG/AMP guidelines provide a map of how to assess pathogenicity, the assessment process is tedious and time-consuming because the relevant data needs to be analyzed from multiple resources (Liu et al., 2019).

In recent years, several computational tools were created to gather all the necessary information to assign the ACMG/AMP evidence codes to each variant to streamline the process (Li and Wang, 2017; Bertoldi et al., 2017; Scott et al. 2019; Xavier et al. 2019; Kopanos et al. 2019; Bouzinier et al., 2022). However, the accuracy of these computational tools is questioned due to the absence of strict criteria for assigning ACMG/AMP evidence codes, infrequent updates of public databases, and missing data points such as segregation and functional studies information (Strande et al., 2018; Niehaus et al., 2020; Basel-Salmon and Sukenik-Halevy, 2022). Moreover, these tools face significant challenges with VUS variants, as the data retrieved for these variants is often scarce or inconsistent (Donohue et al., 2021). Thus, more accurate tools are needed to enable geneticists to interpret variants reliably.

Another important aspect in correctly identifying the causative variants relies on the reference transcript. It is common practice to use biologically supported sequences, such as the MANE Select transcripts (Morales et al., 2022), as the reference transcript in clinical variant interpretation. However, it is not always sufficient to use the effect of a variant on reference transcripts, and the effect of the variant should be analyzed in other clinically relevant transcripts, such as the MANE Plus Clinical set, which includes transcripts where MANE Select transcripts are not enough to report all of the P or LP variants that are present in public resources. Although MANE Select and MANE Plus Clinical transcript sets provide the clinician with most of the relevant transcripts, there are still cases in which the effect of a variant is established as P or LP on an alternative isoform that is not present in the MANE Plus Clinical set.

Another significant issue in variant interpretation is that automated computational tools assess only the pathogenicity of a specific variant without incorporating phenotypic data or a preliminary diagnosis (Ackerman et al., 2016; Berrios et al., 2021). Given the rapid increase in both the number and the scope of NGS-based diagnostic tests each year, and the limited availability of trained geneticists to analyze these data, multiple new approaches have emerged (De La Vega et al., 2021; Jacobsen et al., 2022; Kelly et al., 2022; Nicora et al., 2022; Tosco-Herrera et al., 2022; Yuan et al., 2022; Meng et al., 2023; Zucca et al., 2024). Here, we introduce Genomize-SEQ, a secondary and tertiary clinical bioinformatics platform that aggregates variant-related information from multiple databases and incorporates it for an enhanced, automated ACMG pathogenicity assessment. This integrated approach enables users to quickly and reliably access all essential variant details from a single interface. The automated ACMG pathogenicity assessment of Genomize-SEQ is more concordant with the ClinGen curators compared to other similar publicly available tools such as Franklin (https://franklin.genoox.com) and VarSome (Kopanos et al., 2019). Moreover, Genomize-SEQ is leveraged with a variant prioritization feature in which variants are categorized according to their relevance to the phenotype or preliminary diagnosis entered by users. Finally, Genomize-SEQ shows the effect of each variant not only on reference transcripts but also on alternative transcripts, allowing clinicians to find clinically relevant variants that are important due to their effect on the alternative transcripts. Thus, Genomize-SEQ overall provides a complete solution for comprehensive variant interpretation to achieve fast and reliable diagnosis in diagnosing rare diseases from NGS data.

## Materials and Methods

### The ClinGen Curated Variant Dataset

The ClinGen Expert Panel manually curated variant dataset was downloaded from the ClinGen repository on 13.02.2024 as a CSV file and converted to VCF format (http://erepo.clinicalgenome.org/evrepo/api/classifications/all?format=tabbed).

Variant annotation was performed on the Genomize-SEQ platform (v8.2.0) using RefSeq (v100) and Ensembl (v108) as annotation sources, and the VEP tool (v108.2). The following databases were also used: ClinVar (11.01.2024) and gnomAD (v3.1.2).

In parallel, the same VCF files were analyzed using VarSome (on 15.02.2024) and Franklin-Genoox (on 30.07.2024).

### Real-World Patient Dataset

The cohort consisting of the data from 215 whole exome sequencing samples was previously resolved by the Kocaeli University Medical Genetics team using the Genomize-SEQ platform. Diagnostic variants were reported based on the recommendations of the ACMG guidelines and subsequent updates (Richards et al., 2015; Abou Tayoun et al., 2018). Among these 215 samples, a total of 286 variants were reported (4 samples with 3 variants, 63 samples with 2 variants, and 149 samples with 1 reported variant).

### TopX Analysis

For TopX analysis, each VCF file, for the 215 samples, was analyzed four times: once with preliminary diagnosis and clinical phenotypes, once with preliminary diagnosis only, once with clinical phenotypes only, and once with no input. Variants in each sample were sorted according to prioritization, and the ranking of the variants reported independently by the clinicians was used for the analysis. In the analysis with no input, the filter set below was applied separately, and variants were sorted according to ACMG pathogenicity. The ranking of the variants reported independently by the clinician was used for the analysis, and this group was labeled as the ‘No VP.’

Filter set: Morbid geneset + Pathogenicity VUS or higher + Exonic (coding) + frequency in all normal population < 1%.

### Exome Sequencing for WES samples

Genomic DNA was extracted from peripheral blood using EZ1 DNA Blood 200 µL Kit (Qiagen, Hilden, Germany). Sequencing libraries were prepared using the QIAseq Human Exome Kit (Qiagen, Germany) according to the manufacturer’s instructions. Sequencing was performed on the NovaSeq 6000 (Illumina).

### VCF Generation

Raw sequencing reads in FASTQ format were aligned to the hg19 reference genome using BWA-MEM (v0.7.17). PCR duplicates were removed using Picard Tools (v1.120). Indel realignment was performed with GATK (v3.1.1). Variant calling was conducted using FreeBayes (v1.3.4) with an allele fraction threshold of 0.2 to detect single-nucleotide variants (SNVs) and small indels. The final variant call format (VCF) file was generated for downstream genomic analysis, with post-processing mainly focusing on the classification of heterozygous (HET, <0.95) and homozygous (HOM, ≥0.95) variants.

### VCF Annotation for Variant Prioritization

Variant annotation was performed on the Genomize-SEQ platform (v8.5.0) using Ensembl (v87) as the annotation source, the VEP annotation tool (v100.2), and the following databases: ClinVar (25.06.2024) and gnomAD (v2.1.1).

### Annotation of the Phenopacket Data

Phenopacket Store v.0.1.19 a dataset (Danis et al., 2025) was analyzed by using the annotation pipeline of the Genomize-SEQ platform using Ensembl (v87) as the annotation source, the VEP annotation tool (v100.2), and the following databases: ClinVar (18.08.2024) and gnomAD (v2.1.1).

### Variant Prioritization

The Genomize-SEQ platform employs a complex decision-tree algorithm to prioritize genetic variants based on genomic, phenotypic, and clinical relevance. This algorithm ranks variants according to key genomic characteristics, including public allele frequencies, internal Genomize-SEQ frequency, zygosity, mode of inheritance, and molecular consequence.

Variant prioritization was performed using the Genomize-SEQ platform, which implements the Resnik semantic similarity method (Köhler et al., 2009) to assess gene-phenotype associations. Disease associations were obtained from several curated databases (ClinVar, ClinGen, GenCC, and MONDO) to enhance clinical relevance. Each variant underwent a multi-layered evaluation consisting of: (i) ACMG-based pathogenicity classification; (ii) assessment of gene-disease and variant–disease associations; and (iii) literature-based evidence extraction. Finally, prioritized variants were classified into eight classes (IA to V; Figure 2B), spanning high-confidence, disease-associated variants related to the preliminary diagnosis input (Tier IA) to those deemed non-relevant or prevalent in the general population (Tier V).

## Results

### ACMG Pathogenicity Concordance

We analyzed the concordance of pathogenicity predictions made by three different tools — two widely used automated variant classification engines, VarSome and Franklin, and our algorithm, Genomize-SEQ — by using the manually curated ClinGen expert panel dataset. We first assessed the pathogenicity of each variant in a 5-tier pathogenicity scheme (Pathogenic [P], Likely Pathogenic [LP], Variant of Uncertain Significance [VUS], Likely Benign [LB], Benign [B]) using RefSeq as the annotation source. We found that both VarSome and Franklin had higher recall scores for P variants (0.959 and 0.916, respectively), while Genomize-SEQ had a better precision score (Genomize-SEQ: 0.732, VarSome: 0.497, Franklin: 0.552) (Figure 1A and 1C, Supplementary Table 9). F1 scores were relatively similar for P variants.

**Figure 1:**
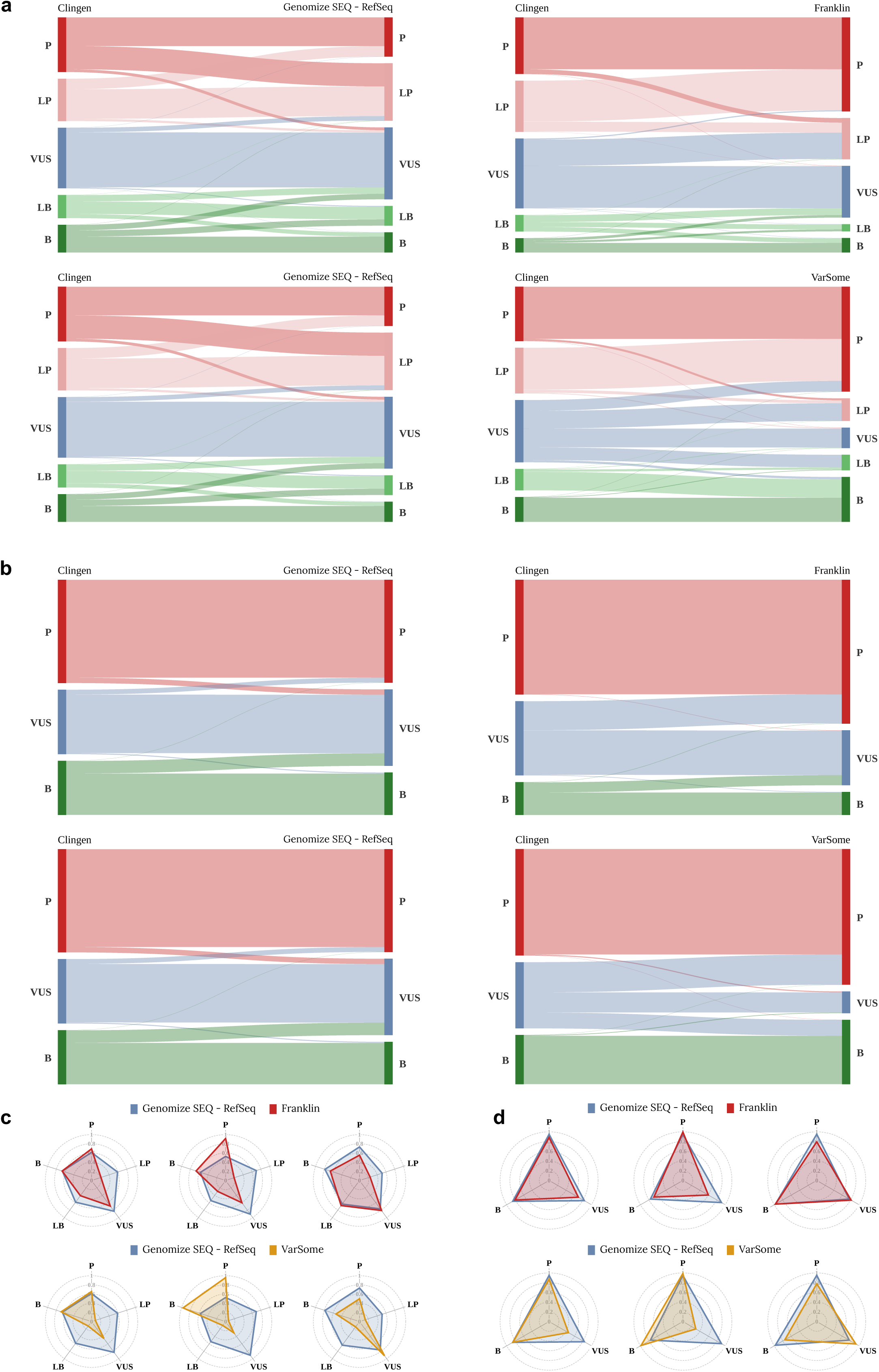
Comparison of automated pathogenicity analysis for variants in the ClinGen expert panel-curated dataset across different variant browsers, using RefSeq as the annotation source. (A-B) Assessment of the pathogenicity of variants in 5-tier classification (A) and 3-tier classification (B). ClinGen expert panel vs Genomize-SEQ (left), ClinGen expert panel vs Franklin (top-right), ClinGen expert panel vs VarSome (bottom-right). (C-D) Radar plot representation of F1-score (left), recall (middle), and precision (right) scores for Genomize-SEQ vs Franklin (top) and Genomize-SEQ vs VarSome (bottom) in 5-tier classification (C) and 3-tier classification (D). Pathogenic [P], Likely Pathogenic [LP], Variant of Uncertain Significance [VUS], Likely Benign [LB], Benign [B].

Next, we analyzed the LP variants and found that 91.82% of the LP variants (1247 out of 1358) were classified as P in VarSome (Supplementary Table 3), while this ratio was 80% (919 out of 1148) in Franklin (Supplementary Table 1), resulting in very low F1, precision, and recall scores for both of these tools (for VarSome F1: 0.087; precision: 0.131; recall: 0.065; for Franklin F1: 0.215; precision: 0.241; recall: 0.194) (Supplementary Table 9). On the other hand, Genomize-SEQ performed better at correctly identifying the LP variants (F1: 0.591; precision: 0.515; recall: 0.699) (Supplementary Table 9).

We then analyzed the VUS variants and found that Franklin again had a tendency to assign higher pathogenicity, with 39.19% of VUS variants (613 out of 1564) classified as P or LP (Supplementary Table 1), yielding an F1 score of 0.689, a precision score of 0.810, and a recall score of 0.600 (Supplementary Table 9). VarSome also had difficulty correctly classifying VUS variants, misclassifying 69.72% of VUS variants (1290 out of 1850) as P, LP, LB, or B (Figure 1A and 1C, Supplementary Table 3), with an F1 score of 0.456, a precision score of 0.920, and a recall score of 0.303 (Supplementary Table 9). On the other hand, Genomize-SEQ was more consistent with the expert panel curation for VUS variants, with an F1 score of 0.830, a precision score of 0.765, and a recall score of 0.908 (Supplementary Table 9).

Next, we analyzed the classification of LB variants and found a different pattern. While VarSome tended to classify LB variants mostly as B (552 out of 636, 86.79%) (Supplementary Table 3), Franklin tended to classify LB variants mainly as B or VUS (257 out of 371, 69.27%) (Supplementary Table 1). On the other hand, Genomize-SEQ was better at correctly classifying LB variants, with an F1 score of 0.586, a precision score of 0.637, and a recall score of 0.543 compared to Franklin and VarSome (for VarSome F1: 0.121; precision: 0.144; recall: 0.105; for Franklin F1: 0.410; precision: 0.677; recall: 0.294) (Supplementary Table 9).

For B variants, the classification accuracy by all three tools was relatively similar, with Genomize-SEQ having the highest precision score (0.786), while VarSome had the highest recall score (0.968) (Supplementary Table 9).

Since the annotation source might be the reason for the discrepancies in the accuracy of estimating pathogenicity, we performed the same analysis using Ensembl as the annotation source and found similar results (Supplementary Figure 1A and 1C, Supplementary Table 2, Supplementary Table 4, Supplementary Table 10).

We also analyzed the pathogenicity estimation by these tools using a 3-tier pathogenicity scheme (Pathogenic [P], Variant of Uncertain Significance [VUS], Benign [B]) and RefSeq as the annotation source. We found that both Franklin and VarSome frequently classified VUS variants incorrectly in the 3-tier scheme (Figure 1B and 1D). Franklin showed a tendency to assign higher pathogenicity, often classifying B variants as VUS or VUS variants as P (Figure 1B, Figure 1D, Supplementary Table 5) with an F1 score of 0.689, precision score of 0.810, and recall score of 0.600 (Supplementary Table 11), while VarSome generally misclassified VUS variants as P or B (Figure 1B, Figure 1D, Supplementary Table 7) with an F1 score of 0.456, precision score of 0.920, and recall score of 0.303 (Supplementary Table 11). On the other hand, Genomize-SEQ outperformed the other tools in terms of F1, precision, and recall scores (Supplementary Table 11), with this difference being particularly evident for VUS variants (F1 score of 0.830, precision score of 0.765, and recall score of 0.908) (Supplementary Table 11). Using Ensembl instead of RefSeq as the annotation source gave similar results (Supplementary Figure 1B, Supplementary Figure 1D, Supplementary Table 12), indicating that the differences in pathogenicity prediction were not due to the annotation source.

These results suggest that, regardless of the annotation source, Genomize-SEQ has superior overall performance compared to VarSome and Franklin for automated pathogenicity prediction. Additionally, Franklin tends to assign higher pathogenicity, whereas VarSome frequently misclassifies VUS variants.

### Decision-tree-based Variant Prioritization

Reliably identifying the relevant variants is as important as accurately assigning their pathogenicity classification in expediting the diagnostic process. To this end, we developed a variant prioritization pipeline that creates a shortlist of variants relevant to the patient’s clinical symptoms. This pipeline integrates the patient’s genomic data with the observed phenotypes and the preliminary diagnosis to evaluate the causative role of each variant (Figure 2A). It then classifies each variant into 3 main and 8 subgroups using information retrieved from over 120 different sources (Figure 2B).

**Figure 2:**
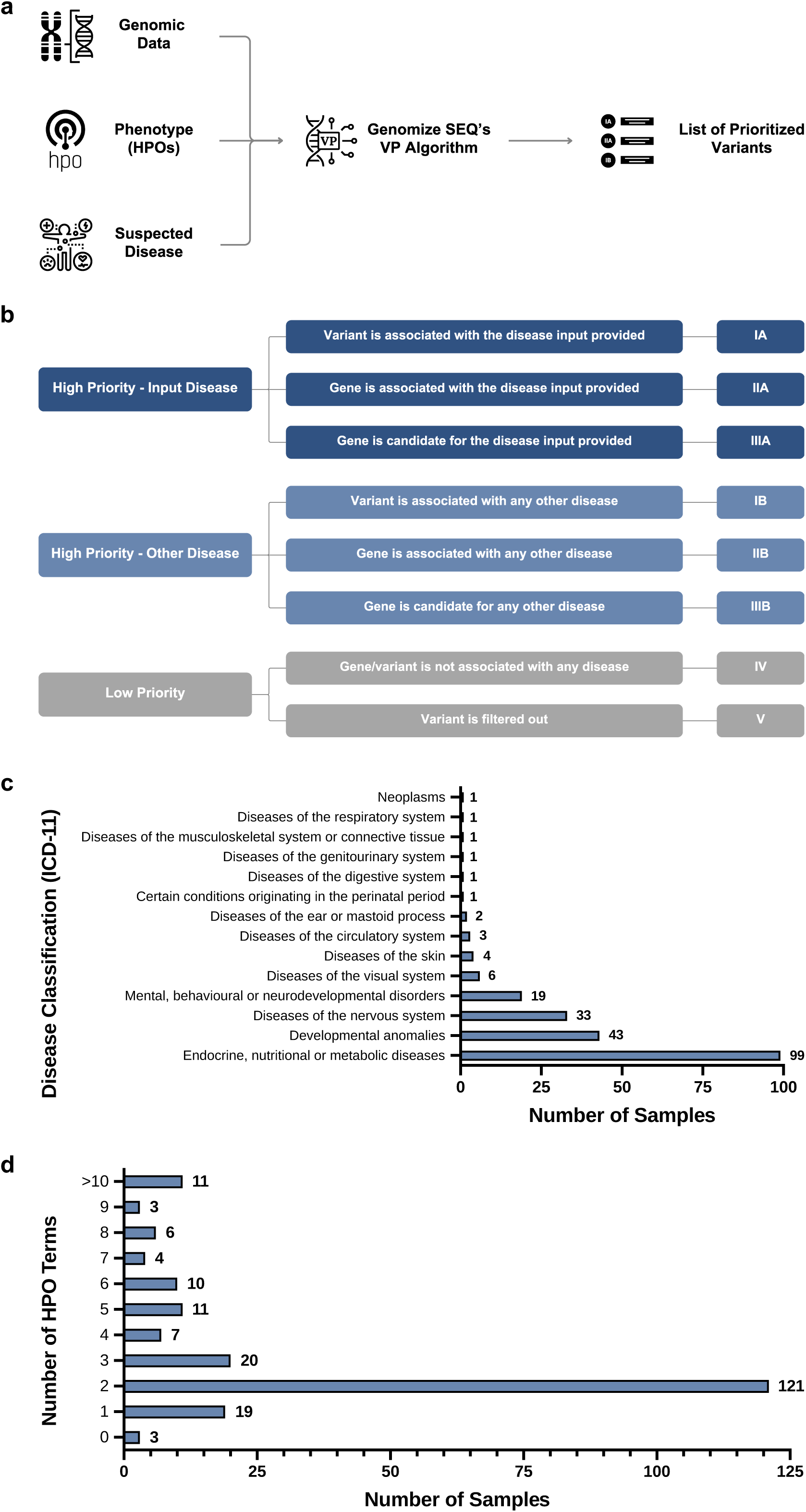
The Variant Prioritization (VP) algorithm in Genomize-SEQ. (A) The VP workflow. (B) Variant tiers in the Genomize-SEQ-VP algorithm. (C) Disease distribution in the sample cohort. (D) Distribution of the number of phenotypes in the sample cohort.

To analyze the accuracy and efficiency of our variant prioritization pipeline, we first performed a case study using a comprehensive real-world patient dataset containing whole exome sequencing (WES) samples from 215 patients who visited the clinic between 2021 and 2023. Their diagnoses span 102 different diseases, including but not limited to inborn errors of metabolism, various neurological diseases, and various developmental disorders, according to the highest-level disease classification of ICD-11 (Figure 2C), with most of the samples representing multiple clinical phenotypes (Figure 2D).

On average, 99.93% of the detected variants in a WES sample were categorized as low-priority variants (classes IV and V), which immediately alleviates the analysis burden and allows the analyst to focus on a small subset of variants (Figure 3A). Out of 286 reported causative variants in 215 cases, 277 (96.85%) were categorized into the high-priority classes (A and B) (Figure 3B and 3C).

**Figure 3:**
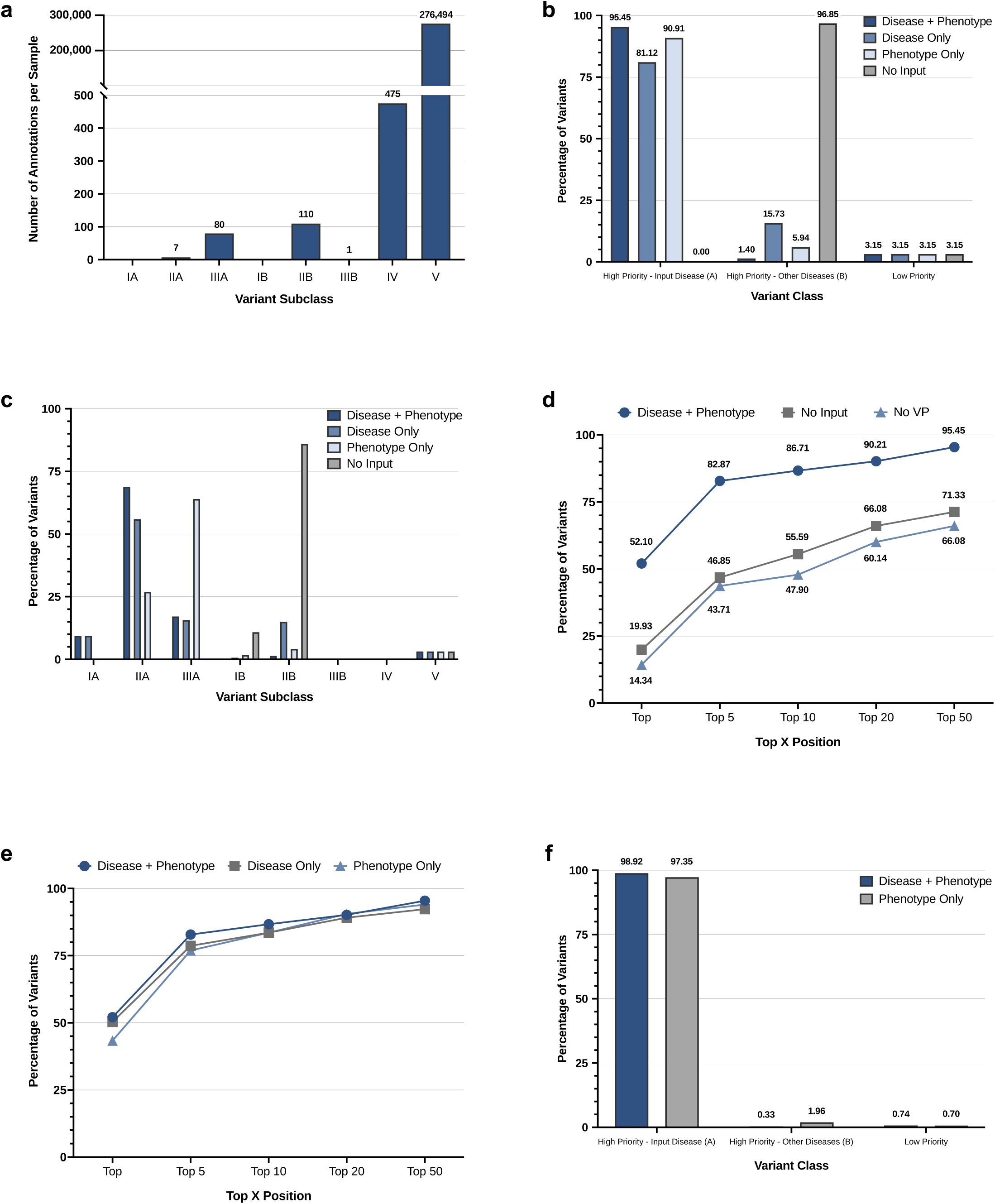
Performance of the Genomize-SEQ-VP algorithm on a dataset of 215 Whole Exome Sequencing (WES) samples. (A) Average number of variants in each VP class per WES sample. (B-C) Percentage of reported causative variants according to the main VP classes (B) and VP subclasses (C). (D) Percentage of cases with reported variants ranked at a certain position (also known as TopX analysis), comparing samples with preliminary diagnosis and phenotype input, no input, and no input with ACMG pathogenicity-based ranking. (E) TopX analysis of reported variants, comparing samples with preliminary diagnosis and phenotype input, preliminary diagnosis input alone, and phenotype input alone. (F) Distribution of variants in the Phenopacket dataset across the main VP classes.

Given the effect of disease and phenotype information on finding the relevant variant, we next analyzed the individual effects of phenotype information and preliminary diagnosis on the classification of variants by our variant prioritization algorithm. When both preliminary diagnosis and phenotype information for the sample are provided, our VP algorithm assigns causative variants as a ‘high priority class-input disease’ variant (A-class) in more than 95% of cases. However, when only preliminary diagnosis or phenotype information is submitted, this ratio drops to 81% and around 91%, respectively (Figure 3B-C). Thus, these results suggest that using both phenotype and preliminary diagnosis information for a sample yields the most optimal result in terms of identifying the causative variants with our variant prioritization algorithm.

Next, we analyzed the effect of our variant prioritization algorithm on the position of the causative variant in the variant list. When preliminary diagnosis and observed phenotypes are provided together, 52% of the reported causative variants were in the first position of the list by our variant prioritization algorithm, and around 90% of the causative variants were in the top 20 positions (Figure 3D-E). Without the clinical information, this ratio dropped to around 20% for the top position and 66% for the top 20 positions (Figure 3D). We then tested whether sorting the variants according to the prioritization provides any benefit, even without phenotype or preliminary diagnosis. To do this, we sorted the variants in the variant list of each sample according to ACMG pathogenicity and applied a commonly used filter set (See Materials and Methods). We found that 14% of the causative variants were at the top of the list, and around 60% of the causative variants were in the top 20 positions for this group which is labeled as ‘No VP’ (Figure 3D). Thus, these results suggest that the VP algorithm ranks the causative variants at the top in most cases, and sorting variants according to their prioritization, even without any phenotype or disease input, ranks causative variants higher compared to applying filters and sorting by ACMG pathogenicity.

Next, we analyzed the effect of phenotype input and preliminary diagnosis input on the ranking of the causative variant separately. We found that providing both phenotype and preliminary diagnosis resulted in improved ranking of the causative variant (Top 1: 52.10%, Top 5: 82.87%, Top 10: 86.71%, Top 20: 90.21%, Top 50: 95.45%) compared to providing only the preliminary diagnosis (Top 1: 50.35%, Top 5: 78.67%, Top 10: 83.57%, Top 20: 89.16%, Top 50: 92.31%) or phenotype information (Top 1: 43.36%, Top 5: 76.92%, Top 10: 83.57%, Top 20: 90.56%, Top 50: 94.06%) (Figure 3E). Including either one or both sets of clinical information resulted in improved variant ranking compared to providing no clinical information (Top 1: 19.93%, Top 5: 46.85%, Top 10: 55.59%, Top 20: 66.08%, Top 50: 71.33%) (Figure 3D vs Figure 3E).

After evaluating our variant prioritization algorithm using real-world data, we further assessed its success by using the recently published Phenopacket Store v.0.1.19, a dataset consisting of case-level, standardized phenotypic information derived from the literature (Danis et al., 2025). We analyzed this dataset for SNVs with our variant prioritization algorithm by entering either the preliminary diagnosis together with the phenotypes (6316 variants) or only the phenotype information (6181 variants). We found that when both preliminary diagnosis and phenotype information are present, our VP algorithm assigns 98.92% of the variants to the ‘high-priority-input disease’ class (A-class), while 0.30% of the variants are assigned to the ‘high-priority-other disease’ class (B-class) (Figure 3F). Similarly, when only phenotype information is used, 97.35% of the variants are classified as the ‘high-priority-input disease’ variant (A-class), and 1.96% of the variants are classified as the ‘high-priority-other disease’ variant (B-class) (Figure 3F).

### Extended Annotation

To address the ‘one variant – many annotations’ issue, a common problem in variant interpretation, we developed a feature called ‘Extended Annotation’ (Figure 4A), which annotates every variant for every transcript in both Ensembl and RefSeq databases, along with their pathogenicities, calculated according to the ACMG guidelines and subsequent updates (Richards et al., 2015; Abou Tayoun et al., 2018). While Extended Annotation increases the total number of annotations by only 1% in an average WES sample, it provides the clinician with established P/LP variants from different resources due to their effect on an alternative isoform. One example of this is the three variants from a previously published paper (Pozo et al., 2022), in which the variant is VUS on the reference transcript but is LP in an alternative transcript that is not in the MANE Plus Clinical set (Figure 4B). Thus, our Extended Annotation feature allows clinicians to find clinically relevant variants that are important due to their effect on alternative transcripts.

**Figure 4:**
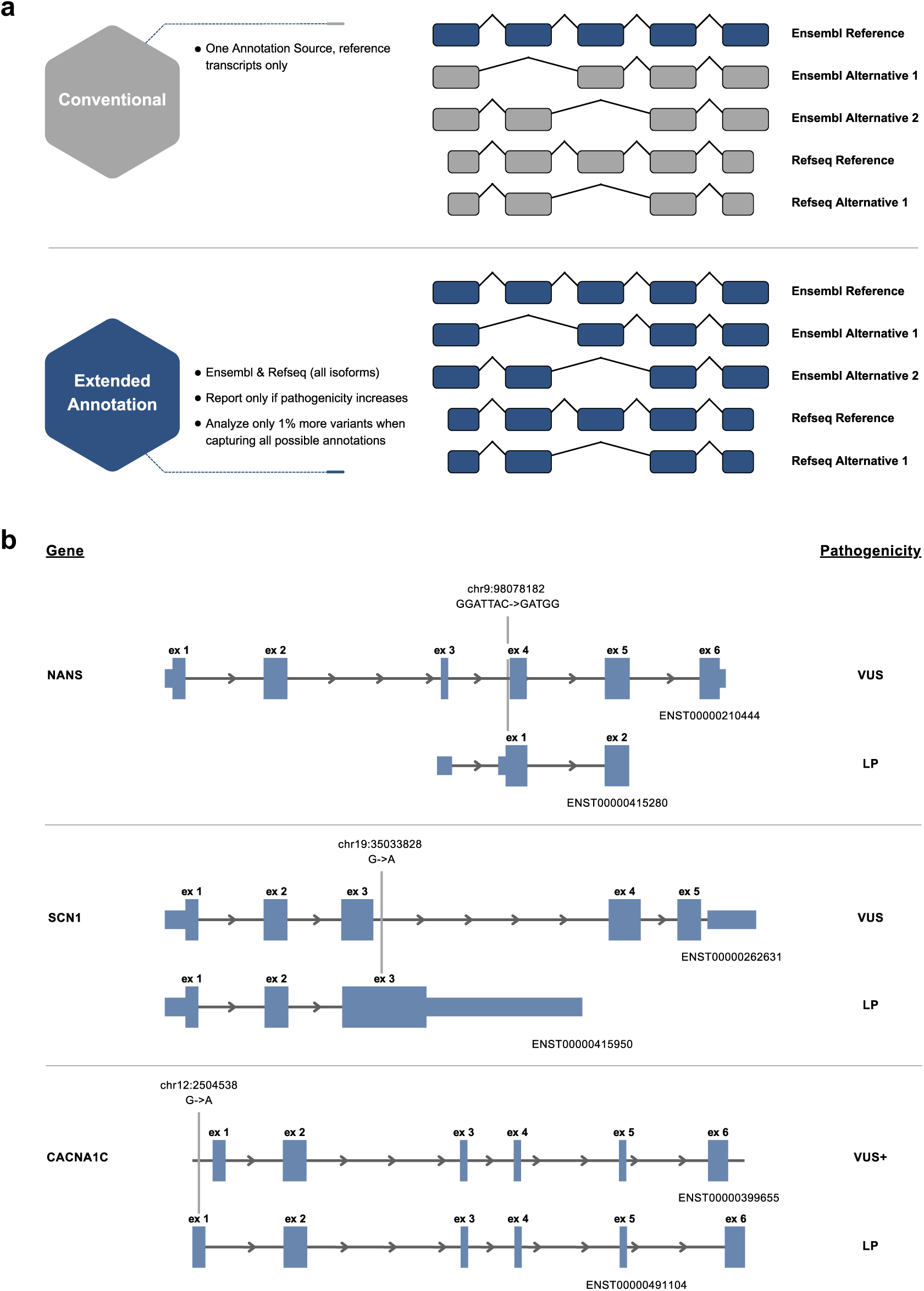
The extended annotation function in the Genomize-SEQ platform. (A) Detailed overview of the extended annotation. (B) Example of pathogenic variants identified by the extended annotation function.

## Discussion

Quickly and reliably identifying disease-causing variants is a primary goal in clinical genomics, especially with the widespread use of next-generation sequencing (NGS) and the significant increase in data from testing. In this study, we present a new tool for geneticists to achieve this goal and demonstrate the functionalities of the software in detail using different datasets.

According to the ACMG guidelines, the pathogenicity of a variant is calculated by assigning 28 different evidence codes, most of which are currently calculated by several commercial software platforms. One issue with the current guidelines is the lack of strict limitations on when to assign certain evidence codes, which can lead to discrepancies between different variant annotation platforms when assessing the pathogenicity of a variant. In this study, using a dataset manually curated by the ClinGen expert panel, we present the first systematic comparison using 6,203 variants as a truth set. The three variant annotation platforms — Genomize-SEQ, Franklin, and VarSome — show significant differences. Among these, Genomize-SEQ had the highest concordance with the expert panel’s assessment, while VarSome and Franklin showed discrepancies compared to the expert panel’s opinion. These discrepancies are especially noticeable for LP, VUS, and LB variants, where both Franklin and VarSome had higher false-negative assignments, as seen in their recall scores (Supplementary Table 9, Supplementary Table 10). In these categories, we observed that Franklin tends to assign higher pathogenicity classes to many variants, while VarSome often misclassifies many VUS variants as B, LB, LP, or P. On the other hand, the main issue for both VarSome and Franklin with P and B class variants is false-positive pathogenicity assignments, which is evident in their precision scores (Supplementary Table 9, Supplementary Table 10). Given the importance of accurate pathogenicity assessment in genetic testing, clinicians should be aware of the tendencies of these software programs in their assessments. Moreover, the primary reason for discrepancies in pathogenicity assessments between these platforms stems from the loose limitations on evidence codes in the current ACMG guidelines. With new ACMG guidelines on the horizon, we believe it would be beneficial for the entire community if the next guidelines imposed stricter rules on pathogenicity assessment to prevent confusion and discrepancies between different platforms.

One important aspect of variant interpretation is to identify the relevant causative variant quickly and reliably. This can be achieved by ranking variants based on the provided disease and phenotype inputs, retrieving information from various databases, including but not limited to gnomAD, MONDO, ClinGen, and ClinVar, a process commonly known as Variant Prioritization (VP) (Cooper and Shendure, 2011; Eilbeck et al., 2017). In this study, we demonstrate the efficacy of our VP algorithm and its success rate using a real-world dataset of 215 WES samples. We found that, on average, 99.93% of the variants in a WES sample are low-priority variants, which significantly reduces the analysis burden on analysts and allows them to focus on a small subset of variants. Furthermore, our VP algorithm achieved a 96.85% success rate in assigning the relevance of reported causative variants in the dataset, further illustrating the efficiency of the algorithm. We also showed that entering a patient’s clinical information significantly improves the ranking of the causative variant compared to samples without any clinical information, thereby simplifying the identification of causative variants. Our analysis indicated that including either prognostic information or clinical phenotypes alone improved variant ranking; however, the best results were obtained by incorporating both when possible. Finally, we assessed the success rate of our VP algorithm using the Phenopacket dataset and observed a 99.3% success rate. In both validation studies, we found that most cases could be resolved within a few minutes. A small minority of cases involved hard-to-diagnose conditions that required more involvement from the analyst. Therefore, our variant prioritization algorithm enables clinicians to identify causative variants accurately and quickly, with minimal risk of overlooking critical genetic variants.

Many different approaches have been developed to achieve efficient variant prioritization using the clinical information provided, with varying levels of success (Javed et al., 2014; Singleton et al., 2014; Smedley et al., 2015; Yang et al., 2015; Bone et al., 2016; De La Vega et al., 2021; Jacobsen et al., 2022; Kelly et al., 2022; Nicora et al., 2022; Tosco-Herrera et al., 2022; Yuan et al., 2022; Meng et al., 2023; Zucca et al., 2024). Although we have demonstrated that our VP algorithm is efficient in prioritizing variants based on case information, a direct comparison with other approaches is not possible at the moment, as the datasets differ among these studies.

The effect of a variant on alternative isoforms is another aspect of clinical variant interpretation that is often overlooked. While most cases can be explained using the GRCh38 genome assembly due to the higher accuracy of canonical transcripts (Pan et al., 2019) and the use of the MANE Plus Clinical transcript set (Morales et al., 2022), there are instances where a case cannot be resolved due to the absence of a relevant transcript in both the reference transcripts and the MANE Plus Clinical set. In this study, we provided an example of such a case and demonstrated how our extended annotation feature can highlight these transcripts for analysts and clinicians. Finally, although the reference transcript set in the GRCh38 assembly is more accurate, many clinical labs continue to use the GRCh37 genome assembly for genetic testing, which lags in terms of transcript accuracy. Thus, this feature further facilitates the diagnostic process for institutions still using GRCh37.

In conclusion, Genomize-SEQ provides fast, reliable, and accurate variant interpretation for geneticists and genome analysts. Recognizing the needs of the community, we created a publicly available search engine that can be accessed at seq.genomize.com/variants or https://variantdb.com. This platform allows geneticists and genome analysts to search for the pathogenicity of human genomic variations by querying gene names, transcript symbols, variant IDs, or HGVS nomenclature (Dunnen et al., 2016), based on the ACMG guidelines and subsequent updates (Abou Tayoun et al., 2018; Richards et al., 2015).

## Supporting information

VarSome annotation result file

Franklin annotation result file

## Data Availability

Annotation result files of VarSome and Franklin can be found as supplementary files. All other relevant data for this study are presented in the main text, and supplemental files are available upon request from the corresponding author.

## Acknowledgements

We would like to thank Devran Karagoz, Mehmet Kaan Demir, Ahmet Can Turkoglu, and the other members of Genomize Bilisim ve Biyoteknoloji Anonim Sirketi for their valuable contributions during the preparation of the manuscript.

## Ethics Declaration

Consent for clinical testing of the 215 samples included permission for the use of anonymized data in research.

---

**Supplementary Figure 1:**
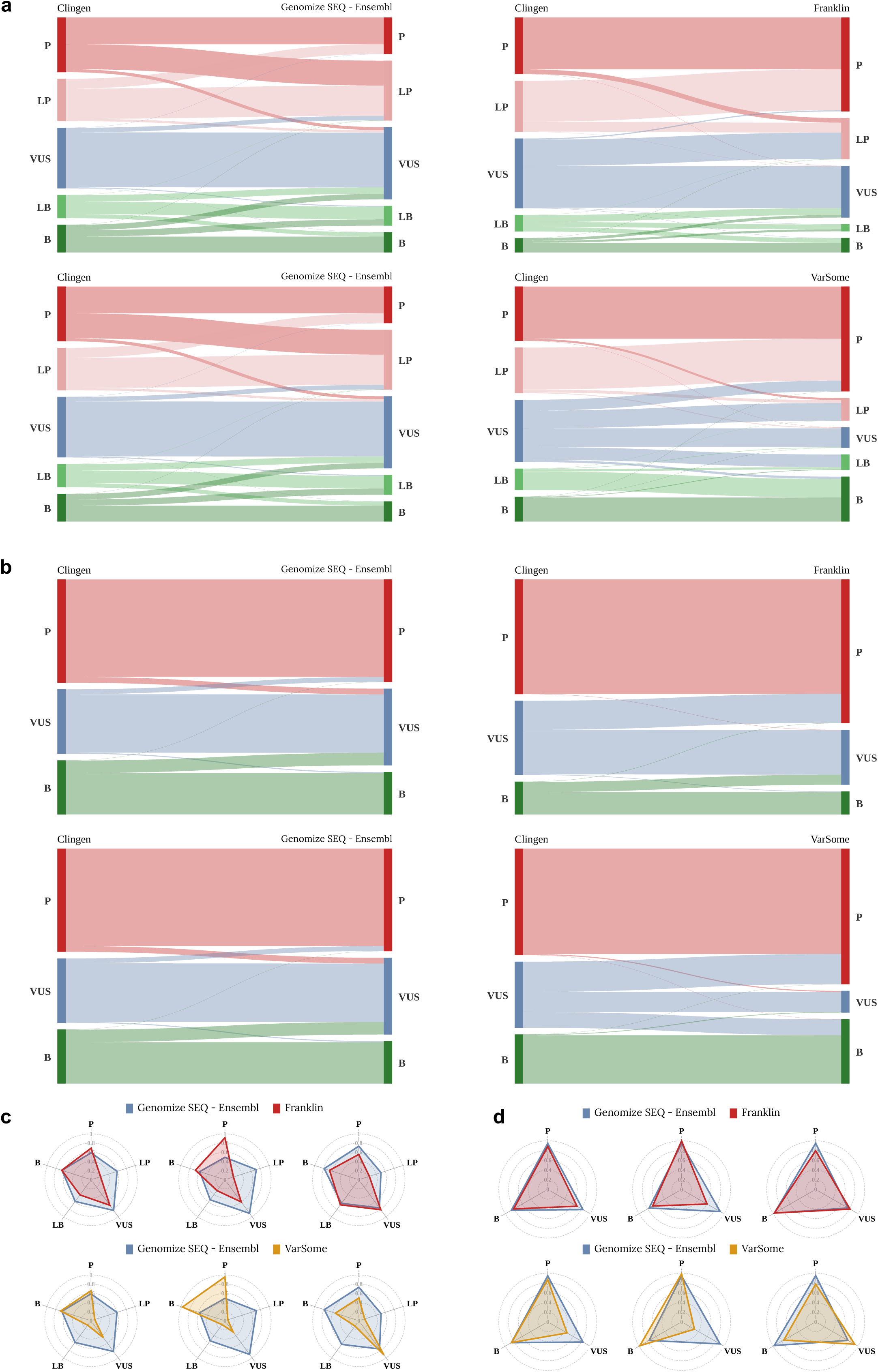
Comparison of automated pathogenicity analysis for variants in the ClinGen expert panel curated dataset across different variant browsers using Ensembl as the annotation source. (A-B) Assessment of the pathogenicity of variants in 5-tier classification (A) and 3-tier classification (B): ClinGen expert panel vs Genomize-SEQ (left), ClinGen expert panel vs Franklin (top-right), ClinGen expert panel vs VarSome (bottom-right). (C-D) Radar plot representation of F1-score (left), recall (middle), and precision (right) scores for Genomize-SEQ vs Franklin (top) and Genomize-SEQ vs VarSome (bottom) in 5-tier classification (C) and 3-tier classification (D). Pathogenic [P], Likely Pathogenic [LP], Variant of Uncertain Significance [VUS], Likely Benign [LB], Benign [B].

