## Supplementary tables for "Genomize-SEQ: An NGS data analysis platform for genomic variant classification and prioritization"

**Supplementary Table 1:** Pathogenicity analysis of the ClinGen Expert Panel manually curated variant dataset in Genomize-SEQ vs. Franklin, using RefSeq as the annotation source in the 5-tier classification system. Franklin is missing a total of 594 variants.

| Genomize-SEQ (RefSeq) | | ClinGen | Franklin | |
|---|---|---|---|---|
| **Count** | **Pathogenicity** | **Pathogenicity** | **Pathogenicity** | **Count** |
| 726 | P | **P** | P | 1163 |
| 585 | LP | **P** | LP | 105 |
| 71 | VUS | **P** | VUS | 1 |
| 265 | P | **LP** | P | 919 |
| 748 | LP | **LP** | LP | 223 |
| 57 | VUS | **LP** | VUS | 6 |
| 1 | P | **VUS** | P | 24 |
| 115 | LP | **VUS** | LP | 589 |
| 1389 | VUS | **VUS** | VUS | 938 |
| 22 | LB | **VUS** | LB | 6 |
| 3 | B | **VUS** | B | 7 |
| 2 | LP | **LB** | LP | 5 |
| 160 | VUS | **LB** | VUS | 155 |
| 318 | LB | **LB** | LB | 109 |
| 106 | B | **LB** | B | 102 |
| 2 | LP | **B** | LP | 2 |
| 138 | VUS | **B** | VUS | 58 |
| 159 | LB | **B** | LB | 46 |
| 401 | B | **B** | B | 216 |

**Supplementary Table 2:** Pathogenicity analysis of the ClinGen Expert Panel manually curated variant dataset in Genomize-SEQ vs. Franklin, using Ensembl as the annotation source in the 5-tier classification system. Franklin is missing a total of 594 variants.

| **Genomize-SEQ (Ensembl)** | | **ClinGen** | **Franklin** | |
|---|---|---|---|---|
| **Count** | **Pathogenicity** | **Pathogenicity** | **Pathogenicity** | **Count** |
| 677 | P | **P** | P | 1163 |
| 629 | LP | **P** | LP | 105 |
| 77 | VUS | **P** | VUS | 1 |
| 247 | P | **LP** | P | 919 |
| 764 | LP | **LP** | LP | 223 |
| 59 | VUS | **LP** | VUS | 6 |
| 1 | P | **VUS** | P | 24 |
| 115 | LP | **VUS** | LP | 589 |
| 1389 | VUS | **VUS** | VUS | 938 |
| 22 | LB | **VUS** | LB | 6 |
| 3 | B | **VUS** | B | 7 |
| 2 | LP | **LB** | LP | 5 |
| 161 | VUS | **LB** | VUS | 155 |
| 318 | LB | **LB** | LB | 109 |
| 106 | B | **LB** | B | 102 |
| 2 | LP | **B** | LP | 2 |
| 137 | VUS | **B** | VUS | 58 |
| 159 | LB | **B** | LB | 46 |
| 401 | B | **B** | B | 216 |

**Supplementary Table 3:** Pathogenicity analysis of the ClinGen Expert Panel manually curated variant dataset in Genomize-SEQ vs. VarSome, using RefSeq as the annotation source in the 5-tier classification system.

| Genomize-SEQ (RefSeq) | | ClinGen | VarSome | |
|---|---|---|---|---|
| **Count** | **Pathogenicity** | **Pathogenicity** | **Pathogenicity** | **Count** |
| 726 | P | **P** | P | 1551 |
| 585 | LP | **P** | LP | 60 |
| 15 | VUS | **P** | VUS | 6 |
| 265 | P | **LP** | P | 1247 |
| 748 | LP | **LP** | LP | 88 |
| 34 | VUS | **LP** | VUS | 23 |
| 0 | P | **VUS** | P | 321 |
| 115 | LP | **VUS** | LP | 525 |
| 1389 | VUS | **VUS** | VUS | 560 |
| 0 | LB | **VUS** | LB | 377 |
| 0 | B | **VUS** | B | 67 |
| 2 | LP | **LB** | LP | 1 |
| 160 | VUS | **LB** | VUS | 16 |
| 318 | LB | **LB** | LB | 67 |
| 106 | B | **LB** | B | 552 |
| 0 | P | **B** | P | 1 |
| 2 | LP | **B** | LP | 0 |
| 138 | VUS | **B** | VUS | 4 |
| 159 | LB | **B** | LB | 19 |
| 401 | B | **B** | B | 716 |

**Supplementary Table 4:** Pathogenicity analysis of the ClinGen Expert Panel manually curated variant dataset in Genomize-SEQ vs. VarSome, using Ensembl as the annotation source in the 5-tier classification system.

| Genomize-SEQ (Ensembl) | | ClinGen | VarSome | |
|---|---|---|---|---|
| **Count** | **Pathogenicity** | **Pathogenicity** | **Pathogenicity** | **Count** |
| 677 | P | **P** | P | 1551 |
| 629 | LP | **P** | LP | 60 |
| 77 | VUS | **P** | VUS | 6 |
| 247 | P | **LP** | P | 1247 |
| 764 | LP | **LP** | LP | 88 |
| 59 | VUS | **LP** | VUS | 23 |
| 1 | P | **VUS** | P | 321 |
| 115 | LP | **VUS** | LP | 525 |
| 1389 | VUS | **VUS** | VUS | 560 |
| 22 | LB | **VUS** | LB | 377 |
| 3 | B | **VUS** | B | 67 |
| 2 | LP | **LB** | LP | 1 |
| 161 | VUS | **LB** | VUS | 16 |
| 318 | LB | **LB** | LB | 67 |
| 106 | B | **LB** | B | 552 |
| 0 | P | **B** | P | 1 |
| 2 | LP | **B** | LP | 0 |
| 137 | VUS | **B** | VUS | 4 |
| 159 | LB | **B** | LB | 19 |
| 401 | B | **B** | B | 716 |

**Supplementary Table 5:** Pathogenicity analysis of the ClinGen Expert Panel manually curated variant dataset in Genomize-SEQ vs. Franklin, using RefSeq as the annotation source in the 3-tier classification system (P/LP shown as P and B/LB shown as B). Franklin is missing a total of 594 variants.

| **Genomize-SEQ (RefSeq)** | | **ClinGen** | **Franklin** | |
|---|---|---|---|---|
| **Count** | **Pathogenicity** | **Pathogenicity** | **Pathogenicity** | **Count** |
| 2324 | P | **P** | P | 2410 |
| 128 | VUS | **P** | VUS | 7 |
| 0 | B | **P** | B | 0 |
| 116 | P | **VUS** | P | 613 |
| 1389 | VUS | **VUS** | VUS | 938 |
| 25 | B | **VUS** | B | 13 |
| 4 | P | **B** | P | 7 |
| 298 | VUS | **B** | VUS | 213 |
| 984 | B | **B** | B | 473 |

**Supplementary Table 6:** Pathogenicity analysis of the ClinGen Expert Panel manually curated variant dataset in Genomize-SEQ vs. Franklin, using Ensembl as the annotation source in the 3-tier classification system (P/LP shown as P and B/LB shown as B). Franklin is missing a total of 594 variants.

| Genomize-SEQ (Ensembl) | | ClinGen | Franklin | |
|---|---|---|---|---|
| **Count** | **Pathogenicity** | **Pathogenicity** | **Pathogenicity** | **Count** |
| 2317 | P | **P** | P | 2410 |
| 136 | VUS | **P** | VUS | 7 |
| 0 | B | **P** | B | 0 |
| 116 | P | **VUS** | P | 613 |
| 1389 | VUS | **VUS** | VUS | 938 |
| 25 | B | **VUS** | B | 13 |
| 4 | P | **B** | P | 7 |
| 298 | VUS | **B** | VUS | 213 |
| 984 | B | **B** | B | 473 |

**Supplementary Table 7:** Pathogenicity analysis of the ClinGen Expert Panel manually curated variant dataset in Genomize-SEQ vs. VarSome, using RefSeq as the annotation source in the 3-tier classification system (P/LP shown as P and B/LB shown as B).

| Genomize-SEQ (RefSeq) | | ClinGen | VarSome | |
|---|---|---|---|---|
| **Count** | **Pathogenicity** | **Pathogenicity** | **Pathogenicity** | **Count** |
| 2324 | P | **P** | P | 2946 |
| 128 | VUS | **P** | VUS | 29 |
| 0 | B | **P** | B | 2 |
| 116 | P | **VUS** | P | 846 |
| 1389 | VUS | **VUS** | VUS | 560 |
| 25 | B | **VUS** | B | 444 |
| 4 | P | **B** | P | 2 |
| 298 | VUS | **B** | VUS | 20 |
| 984 | B | **B** | B | 1354 |

**Supplementary Table 8:** Pathogenicity analysis of the ClinGen Expert Panel manually curated variant dataset in Genomize-SEQ vs. VarSome, using Ensembl as the annotation source in the 3-tier classification system (P/LP shown as P and B/LB shown as B).

| Genomize-SEQ (Ensembl) | | ClinGen | VarSome | |
|---|---|---|---|---|
| **Count** | **Pathogenicity** | **Pathogenicity** | **Pathogenicity** | **Count** |
| 2317 | P | **P** | P | 2946 |
| 136 | VUS | **P** | VUS | 29 |
| 0 | B | **P** | B | 2 |
| 116 | P | **VUS** | P | 846 |
| 1389 | VUS | **VUS** | VUS | 560 |
| 25 | B | **VUS** | B | 444 |
| 4 | P | **B** | P | 2 |
| 298 | VUS | **B** | VUS | 20 |
| 984 | B | **B** | B | 1354 |

**Supplementary Table 9:** Statistical analysis of F1, precision, and recall scores for the pathogenicity analysis in Genomize-SEQ, Franklin, and VarSome, using RefSeq as the annotation source in the 5-tier classification system, based on the ClinGen Expert Panel manually curated variant dataset.

| Pathogenicity | Genomize-SEQ (RefSeq) | | | VarSome | | | Franklin | | |
|---|---|---|---|---|---|---|---|---|---|
| | *F1* | *Precision* | *Recall* | *F1* | *Precision* | *Recall* | *F1* | *Precision* | *Recall* |
| **P** | 0.611 | 0.732 | 0.525 | 0.655 | 0.497 | 0.959 | 0.689 | 0.552 | 0.916 |
| **LP** | 0.593 | 0.515 | 0.699 | 0.087 | 0.131 | 0.065 | 0.215 | 0.241 | 0.194 |
| **VUS** | 0.830 | 0.765 | 0.908 | 0.456 | 0.920 | 0.303 | 0.689 | 0.810 | 0.600 |
| **LB** | 0.586 | 0.637 | 0.543 | 0.121 | 0.144 | 0.105 | 0.410 | 0.677 | 0.294 |
| **B** | 0.663 | 0.786 | 0.573 | 0.690 | 0.536 | 0.968 | 0.668 | 0.665 | 0.671 |

**Supplementary Table 10:** Statistical analysis of F1, precision, and recall scores for the pathogenicity analysis in Genomize-SEQ, Franklin, and VarSome, using Ensembl as the annotation source in the 5-tier classification system, based on the ClinGen Expert Panel manually curated variant dataset.

| Pathogenicity | Genomize-SEQ (Ensembl) | | | VarSome | | | Franklin | | |
|---|---|---|---|---|---|---|---|---|---|
| | *F1* | *Precision* | *Recall* | *F1* | *Precision* | *Recall* | *F1* | *Precision* | *Recall* |
| **P** | 0.587 | 0.732 | 0.490 | 0.655 | 0.497 | 0.959 | 0.689 | 0.552 | 0.916 |
| **LP** | 0.592 | 0.505 | 0.714 | 0.087 | 0.131 | 0.065 | 0.215 | 0.241 | 0.194 |
| **VUS** | 0.829 | 0.762 | 0.908 | 0.456 | 0.920 | 0.303 | 0.689 | 0.810 | 0.600 |
| **LB** | 0.586 | 0.637 | 0.542 | 0.121 | 0.144 | 0.105 | 0.410 | 0.677 | 0.294 |
| **B** | 0.663 | 0.786 | 0.574 | 0.690 | 0.536 | 0.968 | 0.668 | 0.665 | 0.671 |

**Supplementary Table 11:** Statistical analysis of F1, precision, and recall scores for the pathogenicity analysis in Genomize-SEQ, Franklin, and VarSome, using RefSeq as the annotation source in the 3-tier classification system (P/LP shown as P and B/LB shown as B), based on the ClinGen Expert Panel manually curated variant dataset.

| **Pathogenicity** | **Genomize-SEQ (RefSeq)** | | | **VarSome** | | | **Franklin** | | |
|---|---|---|---|---|---|---|---|---|---|
| | *F1* | *Precision* | *Recall* | *F1* | *Precision* | *Recall* | *F1* | *Precision* | *Recall* |
| **P** | 0.949 | 0.951 | 0.948 | 0.87 | 0.776 | 0.99 | 0.885 | 0.795 | 0.997 |
| **VUS** | 0.830 | 0.765 | 0.908 | 0.456 | 0.92 | 0.303 | 0.689 | 0.81 | 0.6 |
| **B** | 0.857 | 0.975 | 0.765 | 0.852 | 0.752 | 0.984 | 0.803 | 0.973 | 0.683 |

**Supplementary Table 12:** Statistical analysis of F1, precision, and recall scores for the pathogenicity analysis in Genomize-SEQ, Franklin, and VarSome, using Ensembl as the annotation source in the 3-tier classification system (P/LP shown as P and B/LB shown as B), based on the ClinGen Expert Panel manually curated variant dataset.

| **Pathogenicity** | **Genomize-SEQ (Ensembl)** | | | **VarSome** | | | **Franklin** | | |
|---|---|---|---|---|---|---|---|---|---|
| | *F1* | *Precision* | *Recall* | *F1* | *Precision* | *Recall* | *F1* | *Precision* | *Recall* |
| **P** | 0.948 | 0.951 | 0.945 | 0.87 | 0.776 | 0.99 | 0.885 | 0.795 | 0.997 |
| **VUS** | 0.829 | 0.762 | 0.908 | 0.456 | 0.92 | 0.303 | 0.689 | 0.81 | 0.6 |
| **B** | 0.857 | 0.975 | 0.765 | 0.852 | 0.752 | 0.984 | 0.803 | 0.973 | 0.683 |
